# Probabilistic forecasting of prefecture-level influenza in Japan accounting for surveillance data release histories

**DOI:** 10.64898/2026.10.09.26364706

**Authors:** Ryutaro Nakagawa, Nami Yamamura

## Abstract

**Background:** Public-health use of influenza surveillance - situational awareness, planning for clinical surge, and issuing advisories or warnings - needs more than an accurate point estimate of next week’s count. What matters operationally is whether a rise is caught early, whether the direction of change is right, and whether an alert threshold is about to be crossed, all with honest uncertainty. For such use a forecast must also respect reporting delay, later data revision, and the calibration of its prediction intervals. Using Japan’s sentinel surveillance, we evaluated short-term probabilistic forecasts that respect the data release history, and we measured what adding other surveillance series contributes, focusing on the properties that matter for public-health decisions.

**Methods:** The target was the number of influenza reports per sentinel site in each of the 47 prefectures. For each forecast we predicted, as quantiles, the value 1-4 weeks after the most recent observed week. We compared three models: a ridge regression trained jointly across prefectures; a gradient-boosting quantile regression (GBQR) that learns from many surveillance series; and their equal-weight combination on the log scale. Six seasons were used to develop the models. The 2025/26 season was held out as a temporal evaluation period after the specification was fixed; because its epidemic profile had already been seen, it is not a fully independent prospective test. We scored forecasts with the weighted interval score (WIS) and checked prediction-interval coverage. For the Japanese target series we used the value first published for each week; some auxiliary series were taken as retrieved and therefore include later revisions.

**Results:** In 2025/26, the combination’s WIS relative to a persistence forecast was 0.841, 0.811, 0.794 and 0.818 at horizons of 1-4 observed weeks - that is, 16-21% lower than persistence. Coverage of the 80% prediction interval was 79.7%, 77.1%, 74.5% and 75.1%, and of the 95% interval 94.3%, 93.4%, 92.6% and 91.8%. Adding auxiliary series improved the GBQR over the development period. However, on the original scale the combination’s edge over ridge regression was not statistically clear, and in some low-level rising phases the combination did worse than ridge.

**Conclusions:** Over the period studied, models that share information across regions scored better than a persistence forecast. For public-health use, however, the operative questions are the real lead time once reporting delay is accounted for, whether rises from low levels and threshold exceedances are caught in time, and whether the upper prediction intervals are wide enough; on these, the present evidence is not yet sufficient. The 2025 change to the sentinel system also means that pre-change advisory and warning thresholds cannot be read mechanically. A prospective evaluation with strictly controlled data availability is needed before such forecasts are used to support advisories or planning.

## 1. Background

Influenza surveillance exists to support public-health action: situational awareness, planning for clinical surge, and the issuing of advisories and warnings. For these purposes the questions a forecast must help answer are practical ones - is activity rising, how fast, and is it about to cross an alert threshold - and they must be answered with the uncertainty made explicit, because a single predicted value cannot convey the risk of a rapid rise or an unexpected turn. Forecasts are therefore better expressed as distributions (quantiles), and judged not only by overall accuracy but by the properties that drive decisions: getting the direction of change right, catching a rise from low levels early, and flagging threshold exceedance. As an overall summary we use the weighted interval score (WIS), which weighs interval width together with the distance from the observed value (Bracher et al., 2021), and we read it alongside these decision-relevant measures.

Regional probabilistic forecasting and the sharing of information across regions are not new. In Japan, Murayama, Shimizu, et al. (2021) forecast regional influenza with uncertainty estimates using commuting data. In the United States, Thivierge et al. (2025) studied how much spatial information helps state-level forecasts. In England, Mellor et al. (2023) forecast hospital admissions over a regional hierarchy and used the system in the 2022/23 season. We therefore do not claim regional forecasting or the use of prediction intervals as the novelty of this study.

A less-examined issue is that the data available when a forecast is issued may differ from the data retrieved later. Reports arrive with delay and are sometimes revised, so an evaluation built on later-curated data can quietly assume information that was not yet available. Forecasting that allows for report revision already exists (Osthus et al., 2019). Accordingly, throughout this study we keep four dates distinct: the observed target week, the release date, the retrieval date, and the version of the values used for scoring.

When training examples are scarce, one option is to learn across regions and across surveillance series. Murayama, Wakamiya, and Aramaki (2021) used multi-task learning across several countries, and Flusion (Ray et al., 2025) combined learning from many influenza-related indicators with a model ensemble. Such gains, however, need not carry over to a different country and surveillance system.

This study has two aims. First, to evaluate, across several seasons and from a public-health standpoint, short-term probabilistic forecasts of Japan’s prefecture-level reports per site that respect the data release history - judging them not only by an overall score but by how well they get the direction right, catch a rise from low levels, and flag threshold exceedance. Second, to separate the contribution of adding auxiliary series from the contribution of combining models, and to identify when performance changes with the epidemic level and with the rising or falling phase, since those are the situations in which an advisory would be considered. The target is a surveillance reporting indicator; it is not a direct estimate of true incidence or of hospital demand.

## 2. Methods

### 2.1 Study design

This is a retrospective evaluation in which we reconstructed the data conditions that held in the past. Six seasons were used for development: 2017/18, 2018/19, 2019/20, 2022/23, 2023/24, and 2024/25. Because the model was refined while looking at the 2023/24 and 2024/25 results, development-period performance reflects some model selection.

The 2025/26 season was a temporal evaluation period: the specification was fixed before scoring it. But the season’s onset timing, peak, resurgence, cumulative value, and number of high-level weeks had been seen beforehand. We therefore do not call it a fully unseen test set, independent external validation, or prospective validation; even an equal-weight combination carries selection bias from the earlier choice of components and features.

The 2020/21 and 2021/22 seasons had very little activity and are excluded from the main performance summary. The main analysis is thus limited to seasons with an epidemic; year-round operation and false alarms outside the epidemic period must be assessed separately.

### 2.2 Target data and release history

The target was the number of reports per sentinel site for the 47 prefectures, from the Infectious Disease Weekly Report (IDWR). Collection began at week 1 of 2016, and the dataset was built to keep the value first published for each observed week. Some re-hosted files cannot be verified as identical to what was released at the time, so first-published values cannot be guaranteed for the entire period.

We confirmed the structure of the release history directly in the data. Each IDWR provisional file is a single-week cross-section and does not contain earlier weeks, and across all 559 national influenza weeks exactly one version exists per ISO week (no week had more than one creation date). The provisional series is therefore **not revised after first publication** within the data we hold: the value the model uses at each origin is the only value ever published for that week - a real-time value. A within-source “first-published vs later-revised” comparison for the target series is thus not reconstructable, because no revised version exists. Two revision concerns remain, and we state both as limitations: the separately published annual confirmed report (*kakuteiho*), which we do not hold, and the re-hosting noted above.

The reports-per-site value depends on the number of reports from sentinel institutions and the number of reporting sites. It is shaped by which institutions take part, by care-seeking, and by diagnosis, testing, and reporting practices, and it is not a per-capita incidence. The set of reporting sentinel sites was changed at week 15 of 2025. Dividing total reports by reports-per-site to recover the number of sites, the national count was about **4,722 just before the change (week ending 2025-03-31) and about 3,883 just after (2025-04-07)** (pre-change median 4,934; post-change median 3,806), consistent with the size of the change. JIHS (2025) has noted concerns about the continuity of health-center-level data, so the effect of the change must be considered even for prefecture-level series.

We detected weeks with a large drop in the number of reporting sites and excluded them from training and evaluation, which separates the structural 2025 change from transient reporting gaps.

### 2.3 Prediction target and the meaning of “weeks ahead”

Let *d* be the issue date, *t* the most recent observed week of the target series available on that day, and the target the reports-per-site value of observed week *t + h*, with *h* = 1, 2, 3, 4. Under this definition, “*h* weeks ahead” is counted from the latest observed week, not from the issue date.

We measured the real lead time directly. Taking each week’s creation date as the issue date, that date falls a median of 3 days after the week ends (range 3-10), i.e. about 9 days after the week begins. Across the 364 evaluation origins, at the moment of issue the *h* = 1 target week is **still in progress in 98% of cases** (it began about 2 days before issue and ends about 4 days after; it had already ended in only 2%). The *h* = 2, 3, 4 target weeks are effectively always in the future, starting a median of about 5, 12, and 19 days after issue; the median number of days until the target week *ends* is about 4, 11, 18, and 25 for *h* = 1-4. That makes *h* = 1 a nowcast of the week in progress, and *h* >= 2 genuine forecasts of future weeks. Because the horizon labels do not line up with the calendar, we report horizons by observed week (*h* = 1-4) alongside the real lead in days, and we read the shortest horizon as an in-progress nowcast rather than a past week. (The creation date is the file’s compilation date and may differ by a few days from actual online availability; the prospective records for 2026/27 carry issue timestamps so this can be checked directly.)

### 2.4 Models and comparators

The primary comparator was a persistence forecast. Its central prediction is the most recent observed value, and its prediction intervals are built from past forecast errors, with a correction that depends on the current level and recent trend. We therefore treat it as a fully specified probabilistic model, not a naive baseline.

The ridge regression pooled the 47 prefectures and learned the log change for each horizon. Its covariates were the prefecture’s level, its recent change, national and neighboring-region trends, and seasonality and holiday indicators. Pooling gives it more training examples than per-prefecture models, at the cost of not fully capturing each prefecture’s own observation process.

The GBQR pooled examples from different regions and surveillance series. The combination model is the equal-weight average of the ridge and GBQR predictions on the log scale. To tell apart the gain from combining models and the gain from adding auxiliary data, we also scored each component on its own.

Further comparators were a per-prefecture autoregression, a seasonal average, an average of seasonal changes, and a simple combination of baseline methods. All model comparisons used the same prefecture-origin-horizon examples.

### 2.5 Auxiliary series and information availability

For GBQR training we used 77 auxiliary series, including US ILINet, WHO FluNet, European and other national surveillance, and additional Japanese surveillance. These series measure different things in different units (ILI, test positivity, hospitalizations). We did not add them directly to Japan’s reporting level; we used them as extra training examples for the same prediction task.

Unlike the first-published Japanese target series, some auxiliary series were taken as retrieved, with publication delays either recorded or set conservatively. Setting a delay does not undo revisions later applied to past weeks, so analyses that use auxiliary series are not a strict real-time reproduction of every input. A sensitivity analysis that varied the assumed auxiliary delays (recorded delays, a uniform 4-week delay, or none) changed the six-season relative WIS by only −0.002 to +0.016 - within the random-seed variation and not distinguishable. The timing of the auxiliary series therefore does not change the conclusions.

We compared four configurations: Japan’s target data only; plus overseas ILI; plus all auxiliary series; and a 38-series set chosen for openness (fewer redistribution constraints). We also separated adding influenza-related indicators from learning across non-influenza diseases.

### 2.6 Training and leakage checks

At each forecast origin we retrained the model using only examples whose ground truth was available by that date. We verified that overwriting future values does not change the forecast at earlier origins, and confirmed this again for the 2025/26 origins (maximum difference 0 for both the linear and the tree components). This check catches only computational dependence on the altered values; it does not rule out revisions already baked into retrieved past series, nor the selection effect of having seen the evaluation season.

### 2.7 Scoring and uncertainty

We scored the forecast distributions with WIS, a proper scoring rule that combines the absolute error of the median with interval scores for several central prediction intervals; lower is better (Bracher et al., 2021). Besides the original-scale WIS, we computed WIS on a log(1 + *y*) scale of both observations and predicted quantiles. The two scales weight errors differently and answer different questions, so we do not treat them as two tests of the same thing (Bosse et al., 2023).

The main summary is the relative WIS against persistence. We report the prefecture/week/season weights used in aggregation and whether we take the ratio of aggregated WIS or the mean of per-example ratios. Secondary measures were 80% and 95% prediction-interval coverage, upper- and lower-interval deviation, agreement in the direction of change, and discrimination of threshold exceedance. We also separated overall WIS from performance specifically on epidemic onset from low levels.

For uncertainty we used 4- and 8-week block bootstraps, a Diebold-Mariano-type comparison that accounts for autocorrelation, and season-level paired tests. Because the loss is first aggregated across all 47 prefectures within a week and the weeks are then resampled in blocks, within-week spatial co-movement is preserved; what is not fully captured is autocorrelation beyond the block length and the small number of effective seasons. Given this spatial co-movement, the overlap between horizons, and the model selection during development, we do not rest any conclusion on a single p-value. Results that pool seven seasons are not a confirmatory test, since six of those seasons were used for development.

## 3. Results

### 3.1 Overall performance in 2025/26

In 2025/26 the combination’s relative WIS was 0.841, 0.811, 0.794 and 0.818 at horizons 1-4 (Table 1), i.e. 15.9%, 18.9%, 20.6% and 18.2% below persistence. Ridge regression gave 0.861, 0.850, 0.831 and 0.852 at the same horizons.

**Table 1.**
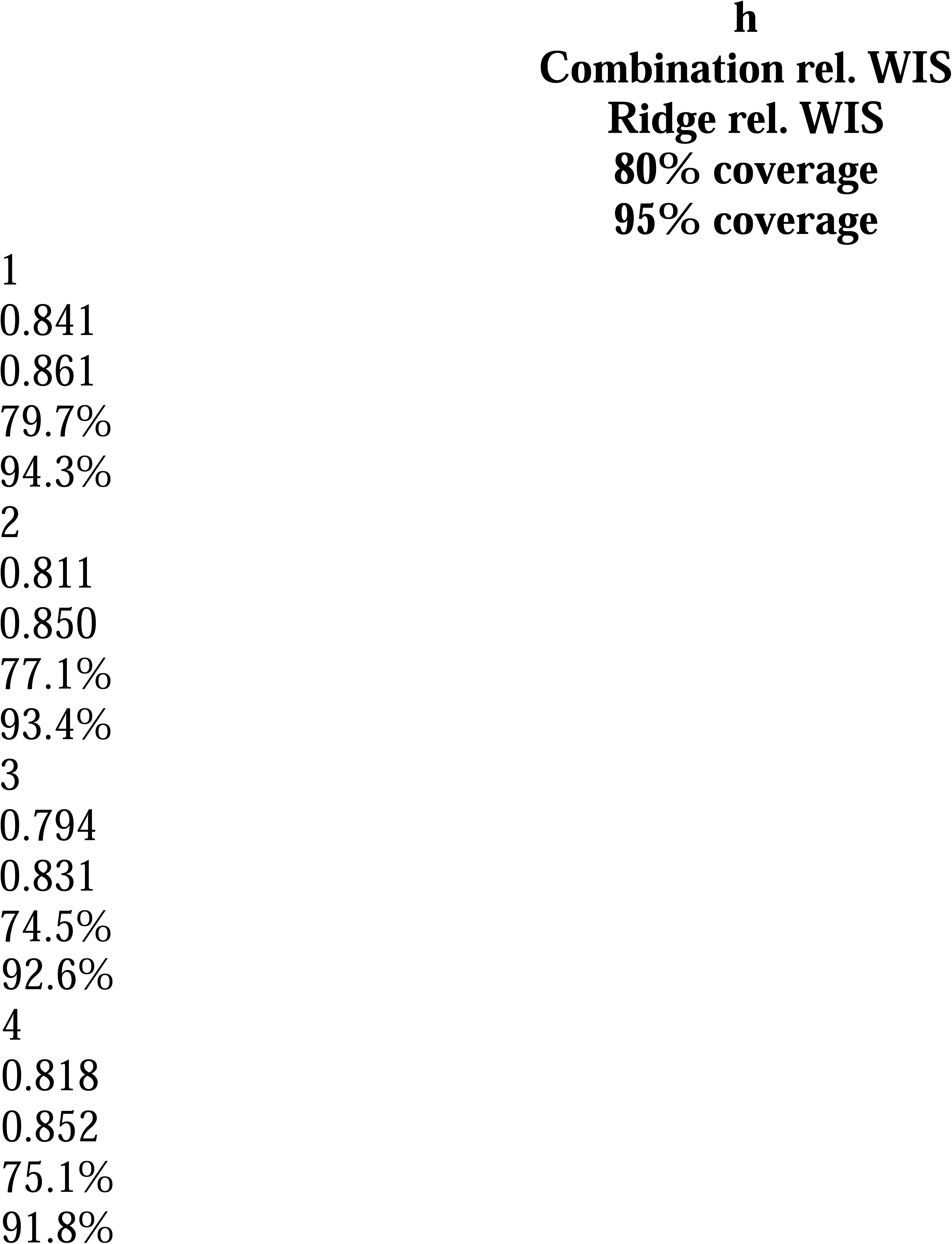
Relative WIS (denominator = persistence) and interval coverage in 2025/26. Horizons are counted from the latest observed week, not the issue date.

The combination beat ridge in point estimates (differences −0.020, −0.039, −0.037, −0.034). However, the 95% intervals all included 0, so an additional advantage on the original scale is not established. This is not evidence of equivalence; it means the observed difference, given the information in the evaluation period, is too small to call.

Across the six development seasons, the combination had a lower relative WIS than persistence in every season, on both scales, at all four horizons. For the seven-season combination-vs-ridge comparison on the log scale, the paired t-test gave p = 0.0081, <0.001, 0.0033 and 0.3909 at *h* = 1-4, and the Wilcoxon signed-rank test gave p = 0.0312, 0.0156, 0.0156 and 0.2969. These are exploratory comparisons that include development seasons and are reported before any multiplicity adjustment.

### 3.2 Contribution of the auxiliary series (development period)

Over the six development seasons, at *h* = 3 (the “next week” horizon) the GBQR relative WIS was 0.847 with Japan’s target data only, 0.821 adding overseas ILI, and 0.796 adding all auxiliary series. With the 38-series open set it was 0.819 - an improvement, but smaller than with the full set.

Adding auxiliary series can therefore help the GBQR. But because the number of training examples, the countries, the indicator types, the observation periods, and the degree of revision all change at once, this does not isolate the causal contribution of any single source, and it is a development-period result with no guarantee of holding in an unseen period.

### 3.3 Interval coverage and phase-specific weaknesses

Interval coverage in 2025/26 tended to fall below nominal at the longer horizons (Table 1). In rapidly rising phases, observations piled up above the upper interval. Stratifying 2025/26 to origins at a level below 10 that were also in a rising phase, the combination had a *higher* (worse) original-scale relative WIS than ridge at all four horizons (Table 2): at *h* = 3, ridge was 0.6834 and the combination 0.8005 (difference +0.117 over 530 examples). All values were still below 1 relative to persistence. The point is that a good overall mean can hide worse performance in a specific phase. Coverage alone cannot say whether the interval shortfall comes from the median under-predicting, from interval width, or from asymmetry.

**Table 2.** Original-scale relative WIS in 2025/26 at origin level < 10 and in a rising phase (difference = combination - ridge; *n* = number of prediction examples, which are not independent). Phases were classified using only information available up to the forecast origin.

| $h$ | Ridge rel. WIS | Combination rel. WIS | Difference | $n$ |
| --- | --- | --- | --- | --- |
| 1 | 0.7476 | 0.7678 | +0.020 | 600 |
| 2 | 0.6848 | 0.7594 | +0.075 | 567 |
| 3 | 0.6834 | 0.8005 | +0.117 | 530 |
| 4 | 0.7475 | 0.8376 | +0.090 | 501 |

### 3.4 Other factors we tried, and a US comparison

We also tried weather, school closures, time-leading of overseas series, joint learning with non-influenza diseases, changes to interval calibration, and correction using early-retrieved series. None gave a consistent improvement. These results depend on how the features were built, on spatial resolution, on publication delay, and on the range of models tuned, and they do not show that any source is useless in general.

As an external check we scored the same approach on US FluSight data (three seasons, 2023/24-2025/26). Our pooled model’s relative WIS to the FluSight baseline was 0.722, which ranked 6th of the 17 methods submitted in all three seasons; it did not reach the FluSight ensemble (0.694) or the top-ranked UMass-flusion (0.650). Our scoring program reproduced the values in Ray et al. (2025) and in the hub’s official report. Our forecasts here were produced retrospectively while the comparators were submitted in real time, so the information conditions differ; we do not read this ranking as evidence of method superiority or of a true cross-country difference.

## 4. Discussion

### 4.1 Main findings and what is new

Over the 2025/26 evaluation period, models that pool learning across prefectures scored better than persistence. But the combination’s margin over ridge was small and not clear on the original scale. For a public-health service, a small gain in an overall score is not by itself a reason to adopt the more complex model: it has to be weighed against the cost of acquiring auxiliary data, of managing release histories, of a more complex implementation, and of keeping all of it running week after week, and against whether the extra complexity actually improves the decisions the service supports.

The contribution here is not a new algorithm. It is the joint study, in Japan’s sentinel surveillance, of release history, cross-region learning, auxiliary series, and phase-specific performance. Handling the release history of the Japanese target series is worthwhile, but because the auxiliary series include revised data, we do not describe the whole study as a strict real-time evaluation.

### 4.2 Learning across different surveillance series

The gain from auxiliary series is consistent with learning timing features of epidemics that recur across different observation systems. It does not mean that overseas epidemics lead Japan’s. Reusing past epidemic shapes as training examples and using current overseas trends as leading indicators are two different hypotheses.

Our study and Ray et al. (2025) differ in prediction target, training period, comparators, and horizon definition. We therefore do not directly compare the size of the relative-WIS reduction to conclude that “the effect is smaller in Japan.” Instead we report the within-study effect and discuss how far it might generalize.

### 4.3 Score gains versus usefulness for decisions

WIS is a useful overall summary, but it does not directly measure the things a public-health user acts on: a missed onset, a false alarm, the lead time left to prepare, or whether an advisory or warning threshold is about to be crossed. We therefore treat the relative-WIS figures as a summary statistic and read them alongside the decision-relevant measures - interval coverage, agreement in the direction of change, and discrimination of threshold exceedance. The scale matters too: on the original scale, errors at high reporting levels dominate, while on the log scale relative change matters more. Doing well on either scale does not guarantee catching a rise from low levels early enough to act on.

Reporting delay sharpens this point. The shortest horizon is partly a nowcast of the not-yet-published value of a week in progress (and, when reports are late, occasionally of a week that has just ended). Such nowcasts aid situational awareness, but the time left to prepare a response is shorter than for a true future forecast. Thresholds set before the 2025 system change should not be applied mechanically to post-change prefecture averages and read as administrative alerts.

### 4.4 Limitations

First, this is a retrospective evaluation with iterative fitting to the development period and prior sight of the evaluation season; the reported intervals and p-values do not capture the uncertainty of the whole model-selection process.

Second, parts of the published versions of the target series, and the revision histories of the auxiliary series, cannot be confirmed, so a complete real-time reproduction cannot be guaranteed.

Third, the 2025 sentinel-system change falls between the training and evaluation periods, and even per-site values can shift if the mix of institutions changes.

Fourth, excluding the non-epidemic seasons means we cannot judge year-round false alarms or behavior at very low activity.

Fifth, prefecture-week observations co-move in space and differ between seasons, so they are not independent replicates; resampling all prefectures together by week keeps within-week spatial dependence, but may still understate the difficulty of generalizing from a few seasons to an unknown one.

Sixth, the exploratory comparison of extra factors involves multiple testing, so the absence of a clear improvement is not proof that a factor is useless.

### 4.5 The validation that comes next

The decisive test is a prospective evaluation that fixes, in advance, the model specification, the rules for deciding what data count as available, and the primary comparisons, and that saves each week’s forecast with an issue timestamp. Persistence and ridge should be saved at the same time, and the evaluation should cover overall scores, interval calibration, lead time measured from the issue date, and threshold exceedance from low levels. A single prospective season would be an important addition, but it would not turn this retrospective study into a prospective one; confirmation across several different epidemic shapes is what is ultimately needed.

## 5. Conclusions

For Japan’s prefecture-level influenza reports per site, models that share information across regions out-scored a persistence forecast over the evaluation period studied, and adding auxiliary series helped the GBQR over the development period; the combination’s extra gain over ridge, however, was not clear on the original scale. From a public-health standpoint, a better overall score is not the same as a service that is safe to act on. The interval shortfall in rapid rising phases, the 2025 sentinel-system change that makes pre-change advisory and warning thresholds no longer directly comparable, reporting delay, and data revision all bear on whether a rise would be flagged in time and without false alarms. These operational questions - early detection, direction, threshold exceedance, and real lead time - should be settled by prospective validation with strictly controlled data availability before such forecasts are used to support advisories or planning, rather than inferred from these retrospective scores.

## Declarations

## Ethics approval and consent

This study used only publicly released, aggregated (prefecture-by-week) surveillance counts, with no individual-level data. It is not human-subjects research, and ethics-committee review and informed consent were not required.

## Author contributions (CRediT)

Ryutaro Nakagawa: conceptualization, methodology, software, formal analysis, data curation, validation, visualization, writing - original draft, writing - review & editing. Nami Yamamura: conceptualization, writing - review & editing.

## Data and code availability

The target data are derived from publicly available sources (IDWR/JIHS and the surveillance series cited in the text). No archived dataset or code DOI is assigned at this time; analysis code and derived datasets are available from the author on reasonable request. Third-party series remain subject to their own terms of use.

## Generative-AI use

The author wrote and approved the manuscript and takes full responsibility for its content. Generative-AI tools (Anthropic Claude, 2026) were used for data processing, model implementation, analysis, internal auditing, literature support, and drafting assistance including English translation. Generative AI is not an author.

---

**Figure A1.**
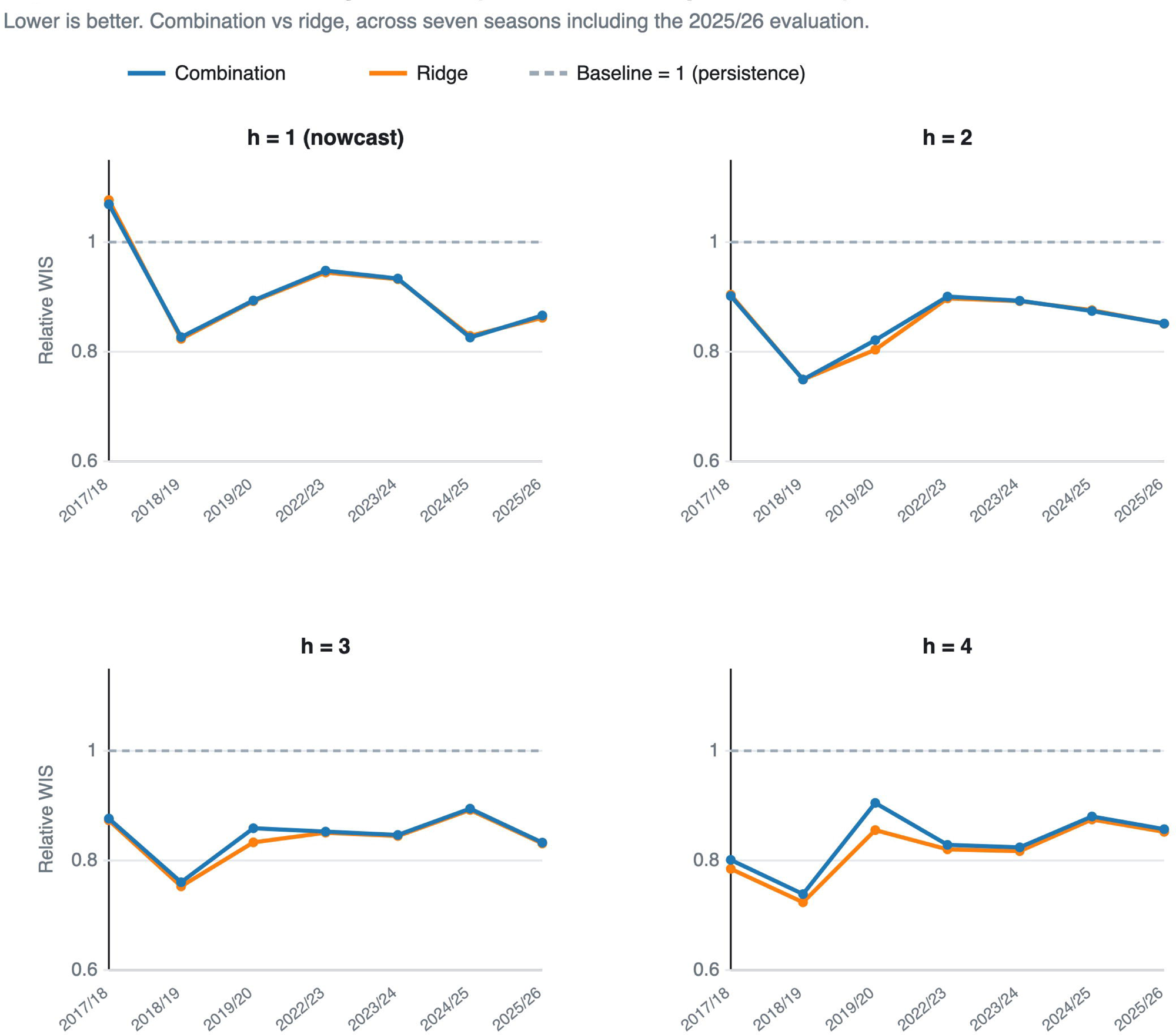
Relative WIS by season (denominator = persistence)

**Figure A2.**
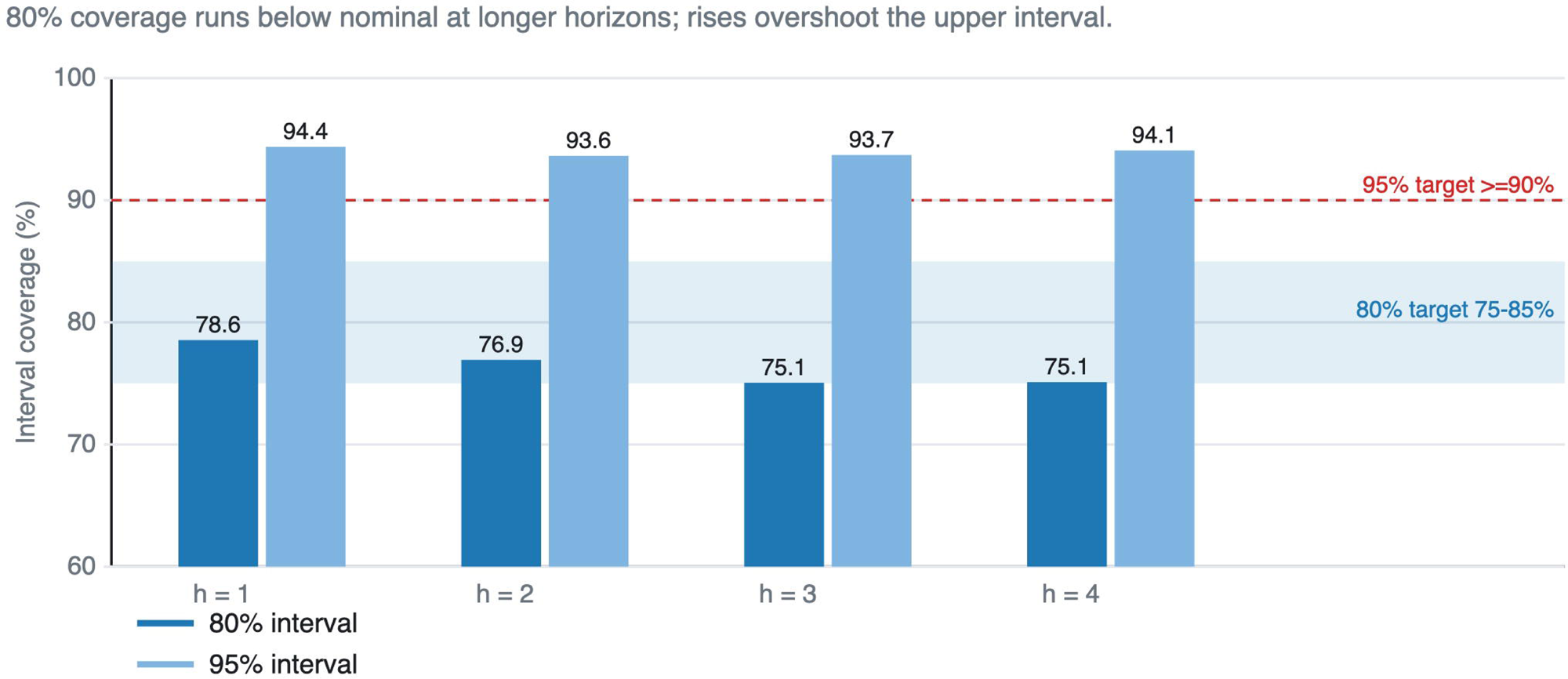
Prediction-interval coverage (2025/26)

**Figure A3.**
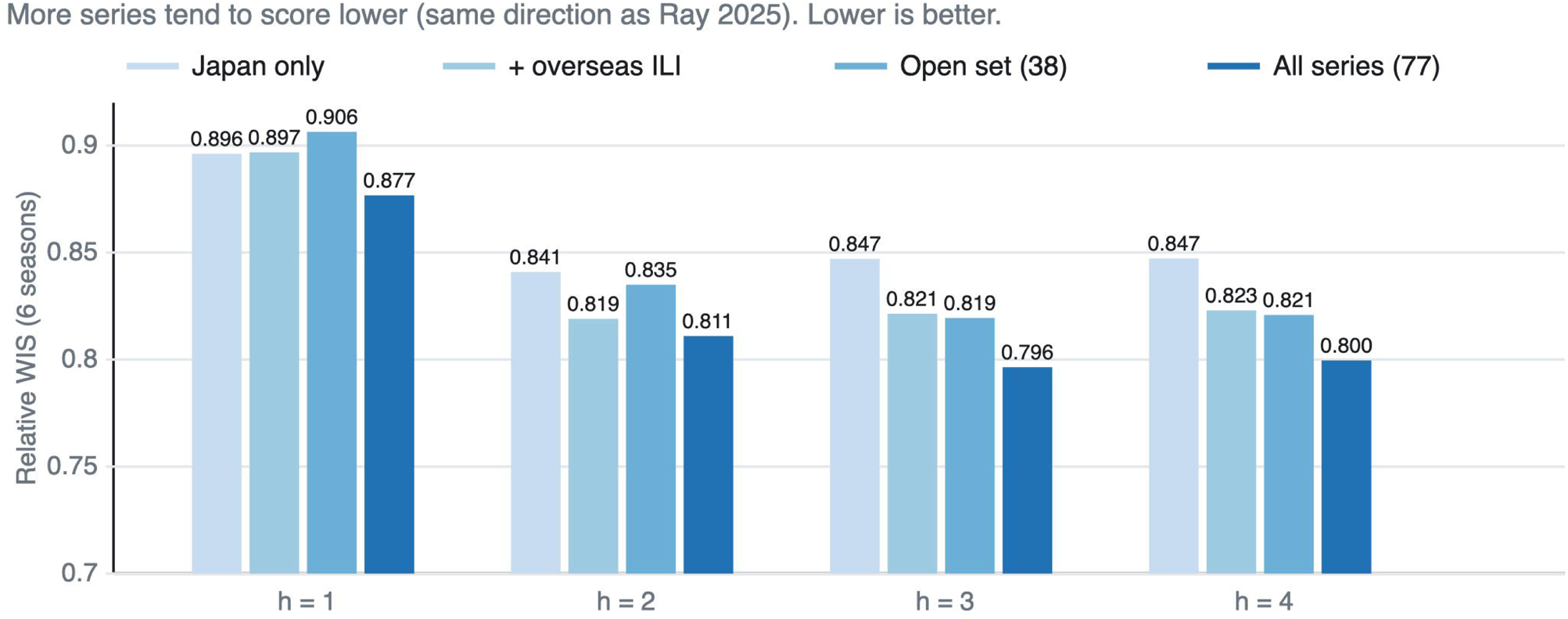
Effect of adding training series (GBQR, 6 seasons)

**Figure A4.**
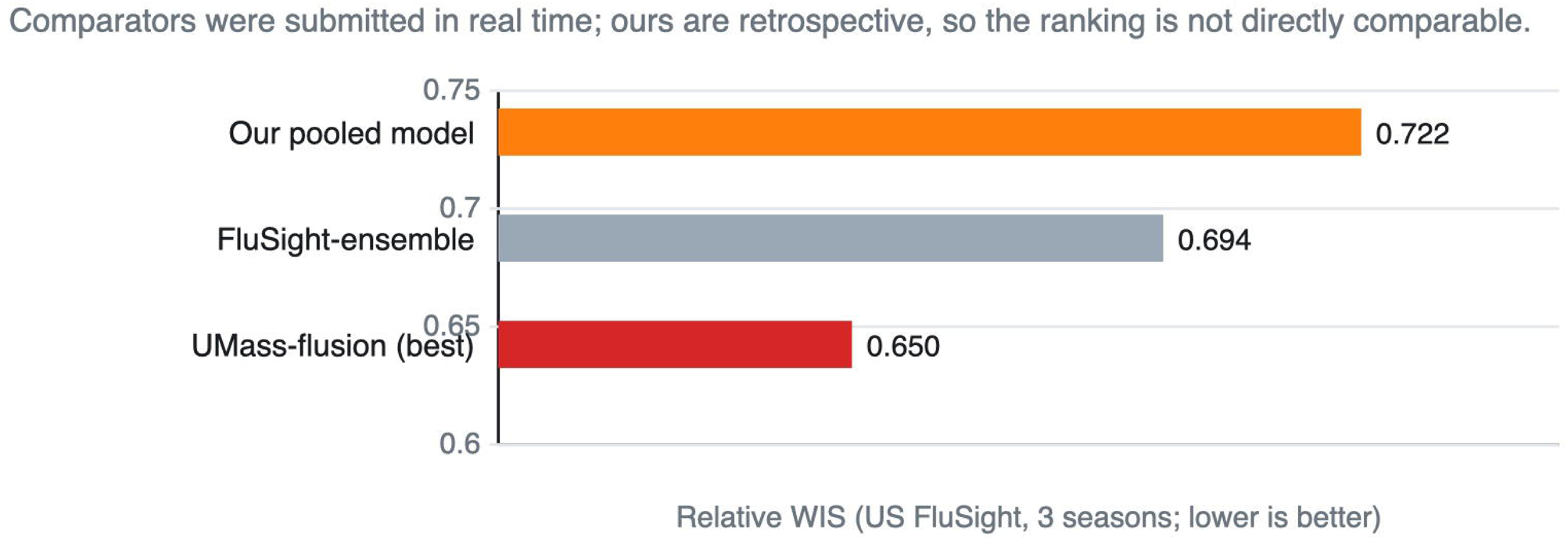
Comparison on US public data (FluSight)

